# Risk assessment of progression to severe conditions for patients with COVID-19 pneumonia: a single-center retrospective study

**DOI:** 10.1101/2020.03.25.20043166

**Authors:** Lijiao Zeng, Jialu Li, Mingfeng Liao, Rui Hua, Pilai Huang, Mingxia Zhang, Youlong Zhang, Qinlang Shi, Zhaohua Xia, Xinzhong Ning, Dandan Liu, Jiu Mo, Ziyuan Zhou, Zigang Li, Yu Fu, Yuhui Liao, Jing Yuan, Lifei Wang, Qing He, Lei Liu, Kun Qiao

**Author notes:** Correspondence should be addressed to Dr. Qiao,; or to Dr. Li,; or to Dr. Liu. These authors contributed equally (Lijiao Zeng, Jialu Li, Mingfeng Liao, Rui Hua).

## Abstract

**Background:** Management of high mortality risk due to significant progression requires prior assessment of time-to-progression. However, few related methods are available for COVID-19 pneumonia.

**Methods:** We retrospectively enrolled 338 adult patients admitted to one hospital between Jan 11, 2020 to Feb 29, 2020. The final follow-up date was March 8, 2020. We compared characteristics between patients with severe and non-severe outcome, and used multivariate survival analyses to assess the risk of progression to severe conditions.

**Results:** A total of 76 (31.9%) patients progressed to severe conditions and 3 (0.9%) died. The mean time from hospital admission to severity onset is 3.7 days. Age, body mass index (BMI), fever symptom on admission, co-existing hypertension or diabetes are associated with severe progression. Compared to non-severe group, the severe group already demonstrated, at an early stage, abnormalities in biomarkers indicating organ function, inflammatory responses, blood oxygen and coagulation function. The cohort is characterized with increasing cumulative incidences of severe progression up to 10 days after admission. Competing risks survival model incorporating CT imaging and baseline information showed an improved performance for predicting severity onset (mean time-dependent AUC = 0.880).

**Conclusions:** Multiple predisposition factors can be utilized to assess the risk of progression to severe conditions at an early stage. Multivariate survival models can reasonably analyze the progression risk based on early-stage CT images that would otherwise be misjudged by artificial analysis.

## Introduction

Since its first report in Wuhan, China, in December 2019, the SARS-CoV-2’s outbreak has quickly became a global public health issue, with a total of 168,019 infected, 6,610 deaths and 148 countries affected as of Mar 16, 2020 (World Health Organization, WHO1). One of main challenges facing its medical care is the lack of effective tools for selecting patients posing a high risk of mortality at an early stage. Previous studies have explored risk factors associated with severity levels of the pneumonia. For example, Zhou et.al2 identified that the older age and a high level of D-dimer are associated with in-hospital death, while Shi et.al3 found that the manifestation of chest CT imaging abnormalities correlates with disease states. However, the joint analysis of time and progression event, as well as the integration of multiple types of input data into risk prediction, have not been thoroughly investigated.

In this study, we reported a retrospectively collected cohort characterized with a large proportion of imported cases. We used this cohort data to develop a competing risks survival model that can predict the real-time risk of progression to severe conditions upon hospital admission for COVID-19 patients.

## Methods

### Study cohort

The cohort includes all adult inpatients admitted at the Third People’s hospital of Shenzhen (Shenzhen, China). The hospital was the only designated center for transfer of COVID-19 patients from other hospitals in Shenzhen city, a metropolis in southern China characterized by a large proportion of migrant workers. Upon hospitalization, all patients were confirmed with SARS-CoV-2 infection but at different stages of the disease. As of Mar 8, 2020, all patients recovered from COVID-19 pneumonia and were discharged from hospital. The study was approved by the Ethical Committee of the Third People’s hospital of Shenzhen. Written informed consent was obtained from each participant patient.

### Data acquisition

The demographical, epidemiological, clinical, laboratory, treatment and progression outcome data were retrospectively collected from electronic medical records, using a standardized form specifically prepared for this study. All authors input, checked and reviewed the data.

### Study definitions

The severe disease conditions were defined based on the Diagnosis and Treatment Protocol For Novel Coronavirus Pneumonia (Trail Version 6) issued by the National Health Commission of China. In brief, a patient can be categorized into severe group if meeting any of the following criteria: 1) respiratory distress (≥ 30 breaths/ min); 2) at-rest oxygen saturation ≤ 93%; 3) arterial partial pressure of oxygen (PaO2)/ fraction of inspired oxygen (FiO2) ≤ 300mmHg (l mmHg=0.133kPa); 4) respiratory failure and requiring mechanical ventilation; 5) shock; 6) with other organ function failure that requires ICU. The non-severe group is defined as meeting any of the following criteria: 1) the clinical symptoms were mild, and there was no sign of pneumonia on imaging; 2) showing fever and respiratory symptoms with radiological findings of pneumonia.

### Statistical analysis

Continuous and categorical variables were presented as mean (standard deviation) and number (%). The between-group significance of difference was evaluated by the one-way ANOVA, chi-square test or Fisher’s exact test. Multiple testing was corrected by Benjamini-Hochberg procedure^1^ to control the false discovery rate (FDR) and to obtain the adjusted p-values. An adjusted p-value smaller than 0.01 was considered as statistically significant. Multivariate logistic regression was used to test the significance of interaction effect whenever possible. All statistical analysis were performed by R (version 3.6.1).

## Results

### Disease process and clinical outcomes

All 338 adult patients (age ≥18) enrolled in this study were treated at the Third People’s hospital of Shenzhen, the only designated hospital for COVID-19 patients in Shenzhen city, China. A typical disease process for the patient can be illustrated in Figure 1. After hospital admission, the patient can either progress to severe conditions, or recover from the pneumonia without any severe progression. Among a total of 76 (31.9%) patients who experienced severity, 18 (5.3%) further progressed to extreme severe or critical conditions. As of Mar 8, 2020, 3 (0.9%) patients died of complications, and 45 (13.3%) remained hospitalized. All others were discharged from hospital with recovered health status as well as sequential negative testing results of SARS-CoV-2. The mean duration from symptom onset to hospital admission is 5.1 days (Supplementary Table 1).

**Figure 1:**
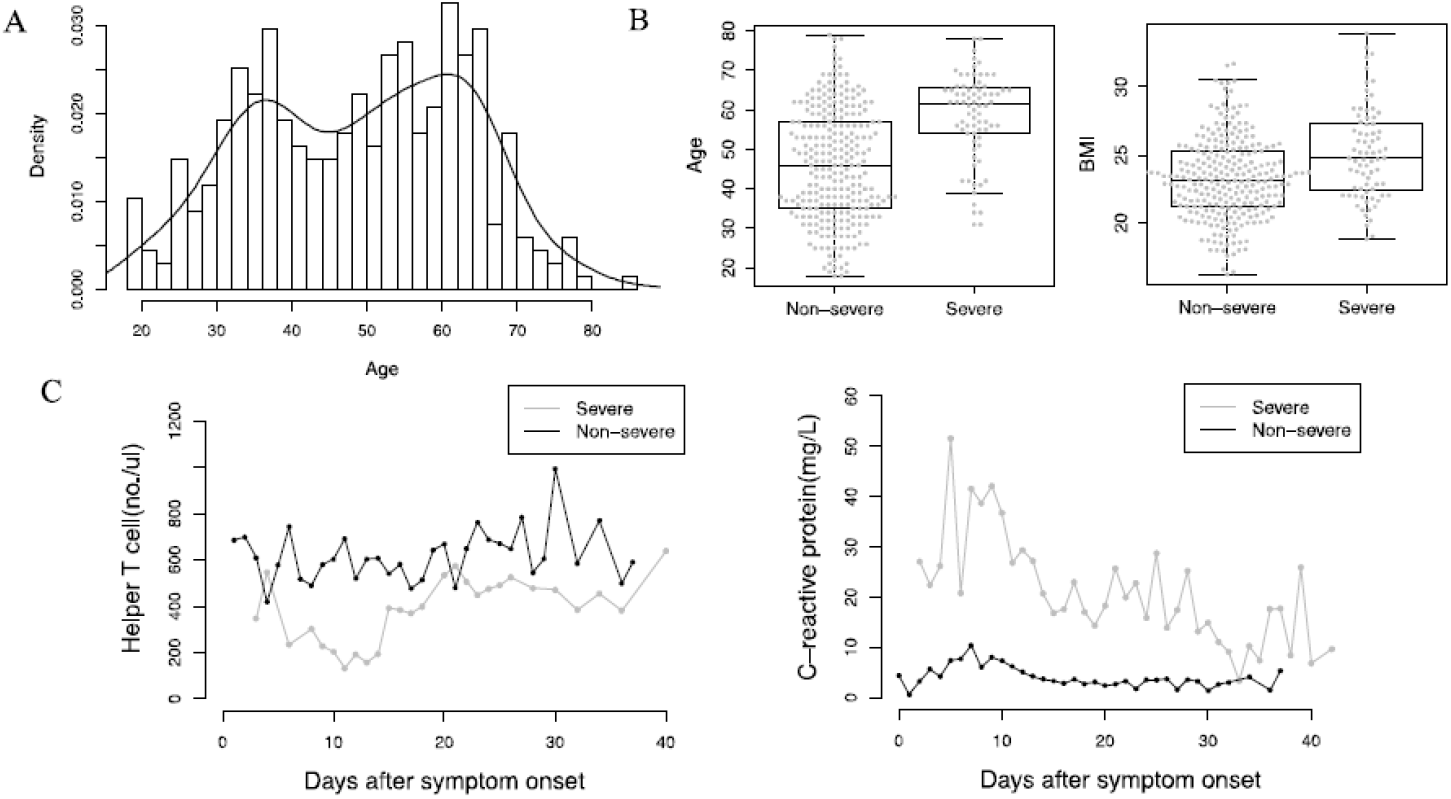
Analysis of representative features significantly associated with the severe group. A, the age distribution of the study cohort, which is overlaid with kernel density estimates (solid curve); B, boxplot summary of the age and BMI as stratified by the severe and non-severe group; C, levels of individual laboratory biomarker plotted along with the time after symptom onset. Data at the same day from multiple patients were collapsed together and only the median value (solid dots) was shown.

The mean time from hospital admission to severity onset is 3.7, to hospital discharge without severe progression is 19.4 days. The mean duration from severity onset to hospital discharge is 21.5 days.

### Characteristics of the study cohort

To evaluate the progression-related features, we classified the patients as severe or non-severe group based on their severity experience during hospitalization. The summary statistics and univariate testing p-values are shown in Table 1. The patient age is bi-modally distributed, with one mode located around 35 years and another peaked at 60 years (Figure 2A). The mean age of the severe group is significantly higher than that of the non-severe group (58.7 versus 46.1). Moreover, the severe group appears to be overweighed as compared to the non-severe group (mean BMI: 25.1 versus 23.2, Figure 2B). The majority of patients (79%) has a travel history to Wuhan region within 14 days before symptom onset. However, exposure history distribution is not significantly different between the two groups. Gender does not have a strong association, nor it has a significant interaction with age under a multivariate logistic regression modeling (data not shown).

**Figure 2:**
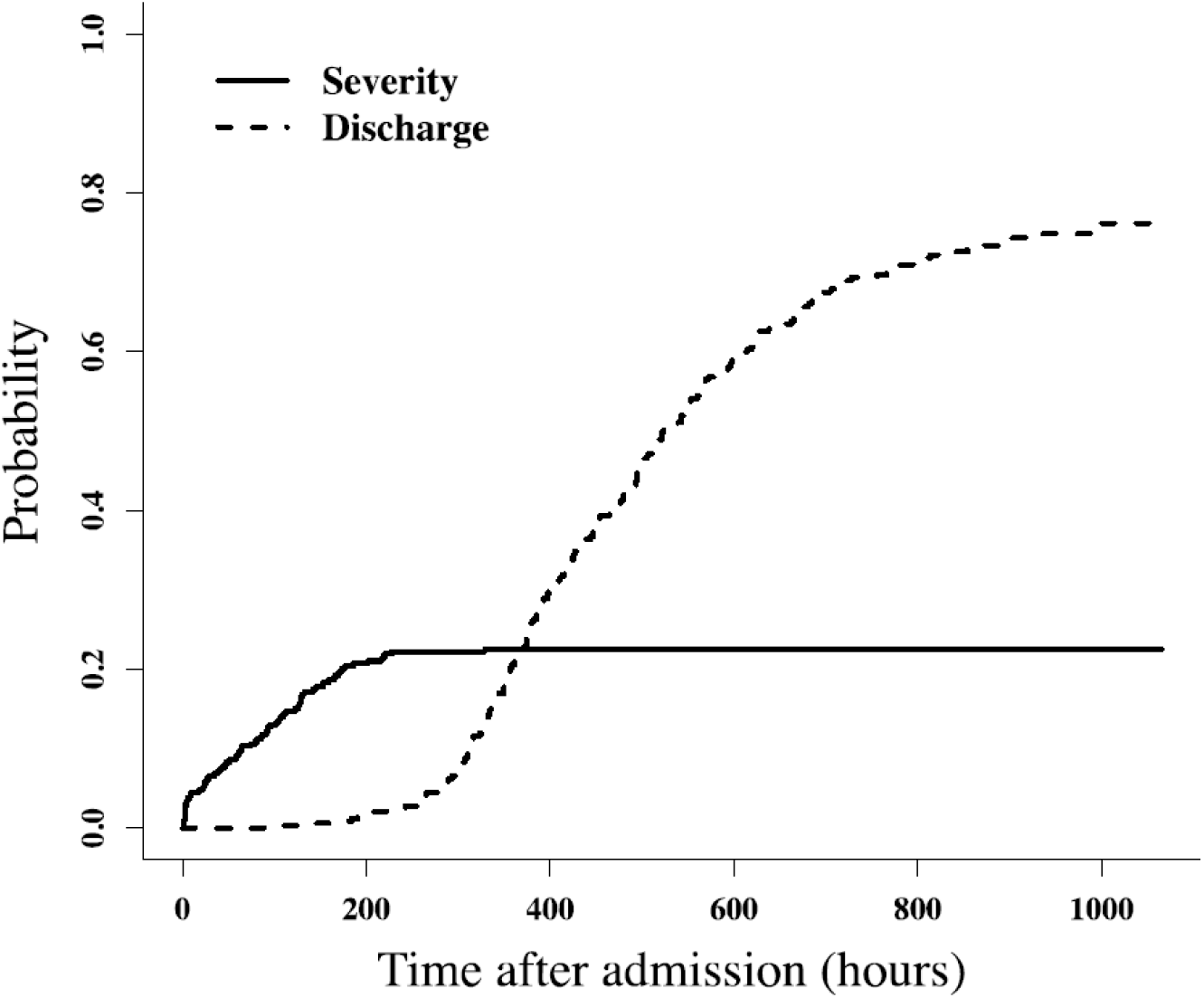
Cumulative probability of developing severe conditions or being discharged from hospital after hospital admission for the study cohort.

Overall, disease history are not significantly enriched in the severe group. However, co-existing cardiovascular or cerebrovascular disease, or co-existing endocrine system disease do have such significance. The common symptoms at disease onset includes fever (64.5%), cough (51.8%), fatigue (13.6%), sore throat (13.0%) and muscle ache (11.8%), while the common ones manifested at hospital admission are fever (60.7%), cough (52.4%), fatigue (15.1%), sore throat (10.9%), muscle ache (10.1%) and diarrhea (9.8%). The severe group has a significantly higher rate of presenting fever when compared to the non-severe group.

### Laboratory testing results

The laboratory testing were performed on patient blood samples collected immediately after hospital admission. The analysis results of their association with severe grouping are summarized in Table 2. The category of significantly associated biomarkers ranges from blood cell counts to kidney function indicators. As for those with an abnormal level of mean measurement, the severe group has elevated levels of Fibrinogen, D-dimer, Lactate dehydrogenase, Glucose, C-reactive protein, Interleukin-6, Erythrocyte sedimentation rate, β2-Microglobulin, and decreased levels of Prealbumin, PaO2/FiO2 ratio, Glomerular filtration rate. Of note, PaO2/FiO2, a ratio used for defining severity onset, already showed a significant difference between the two groups upon admission (Figure 2B). This suggests that disease progression signal is present at an early stage.

To investigate the dynamic variation of blood cell counts, we plotted the counts against the time since symptom onset, stratified by the severity grouping (Figure 2C). The severe group has decreased number of both helper T cells and cytotoxic T cells in the first 10 days after symptom onset. The cell number increased in next 10 days and plateaued after then. Such pattern suggests that the T cell depletion may function as one important mechanism for the progression COVID-19.

### Pattern of disease progression

To assess the risk pattern of disease progression during hospitalization, we performed competing risks analysis on the time-to-event data. The motivation to apply such type of analysis method is that a patient censored because of hospital discharge will certainly not experience severity onset. The risk of severe progression assessed without considering the competition would thus be overestimated because the patients who would never progress were treated as if they could progress. As shown in Figure 3, the cumulative probability (aka. cumulative incidence function, CIF) of severity onset continues to increase immediately after hospital admission, but arrives its change point at about day 10. In contrast, the cumulative incidence of the competing risks event, that is, being discharged because of recovery, is much smaller during the high risk period of severity onset. However, such incidence dramatically increases from around day 12 to 29.

**Figure 3:**
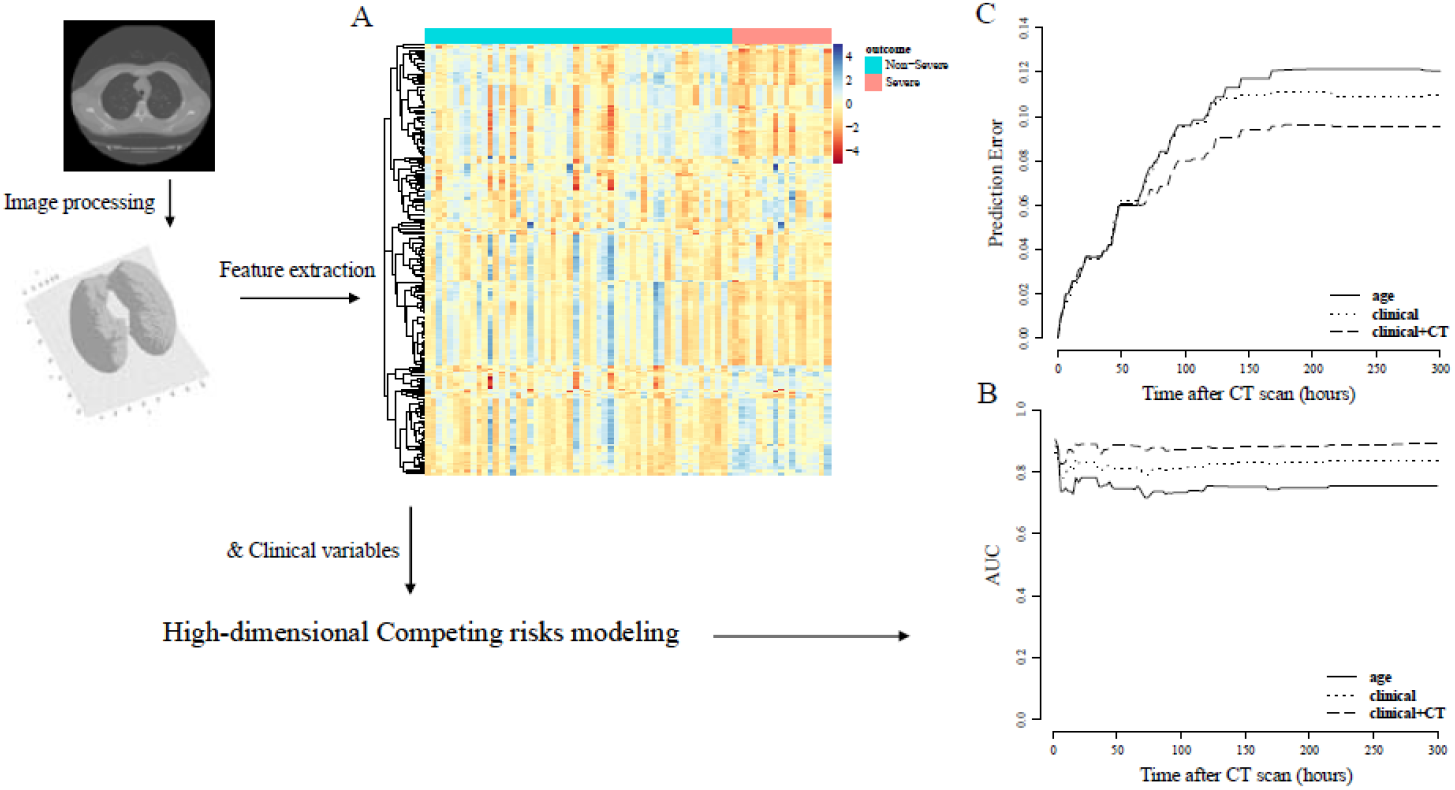
Workflow of the CT-based risk prediction model development. A, heatmap analysis of quantitative features extracted from CT images, stratified by progression outcome; B, model prediction performance evaluation by time-dependent ROC method; C, model prediction performance evaluation by time-dependent prediction error method.

### Risk prediction of progression

To personalize the time-dependent risk assessment of severe progression, we incorporated CT image data with clinical information for the risk modeling (Figure 4). A total of 348 quantitative features were extracted from 3-dimensionally re-constructed chest CT scan images generated upon hospital admission. Clustering analysis showed that there exists a subset of features that can distinguish severe from non-severe group (Figure 4A). We then computed same features from earliest CT scans after admission for each patient, and trained a risk prediction score using high-dimensional competing risk modeling. We used time-dependent ROC and time-dependent prediction error to evaluate the model prediction performance on validation datasets. The model integrating CT and clinical variables significantly outperformed the univariate model and multivariate model using only clinical information (Figure 4B, 4C). The best model achieved a mean AUC of 0.880 (0.011) and a mean prediction error of 0.079 (0.024). We also developed a model integrating clinical variables and laboratory biomarkers tested at a time close to hospital admission. This model has a mean AUC of 0.884 (0.049) and a mean prediction error of 0.103 (0.031) (Supplementary Figure 1).

We presented two cases of study to exemplify the usage of the CT image-based risk assessment tool. Case 1 has ground-glass opacity, while case 2 has no visible abnormality on CT images. We first computed the risk score based on image processing results and then calculated the CIF at each time point within 5 days after the time of CT scan. The cumulative probabilities of developing severity for case 1 within next 1, 3, and 5 days are 0.215, 0.427, 0.613, as compared to 0.001, 0.003, 0.005 for case 2, respectively.

## Discussion

We delineated the characteristics of a retrospective cohort of 338 adult patients collected at a single center from Shenzhen city of China, and developed a non-invasive method to evaluate the risk of progression to severe conditions. The independent predisposition factors of progression include old age, high BMI, fever, and co-existing hypertension or diabetes diseases. However, using age as a single prognostic factor could lead to erroneous results since young patients were not necessarily progression-free. Different from previous studies4-6, the severe group in this cohort has a significantly higher proportion of patients with fever symptom at admission (82.9% vs. 54.2%). Moreover, we identified, for the first time, that overweight is associated with disease severity. These findings benefit the risk assessment analysis as we showed that a model combining these indicators can substantially improve the prediction performance as compared to a model that only contains univariate predictor (mean time-dependent AUC= 0.824 versus 0.751).

SARS-CoV-2 entry host cell through angiotensin-converting enzyme 2 (ACE2)7, whose expression can be enhanced by the usage of hypertension medicine such as ACE inhibitors or angiotensin II type-I receptor blockers (ARBs)8. Whether such medication is a causal factor for a higher risk of COVID-19 progression is thus under investigation9. In our study, we observed that the co-occurred hypertension disorder is significantly related to severe progression, but we did not found association between the medication and severity outcome among hypertension patients.

COVID-19 pneumonia is a multistate disease with clinically relevant intermediate endpoint like severity onset. Most survival data analyses set the onset as the primary end point, and censor recovery or hospital discharge. However, when competing risks of severity onset are present, this analytical method induces bias. In this study, the risk of severe progression assessed without considering the competition would be overestimated because the patients who would never progress (those who discharged from hospital without progression) were treated as if they could progress. The extent of such bias and its adjustment by competing risks modeling have been evaluated in clinical virology and oncology research10-13. We incorporated high-dimensional variable selection techniques into the competing risks modeling so that quantitative image features can be extensively evaluated according to their contribution to risk prediction. Our evaluation results showed that incorporating CT image can significantly improve the prediction performance as compared to those only based on demographical and clinical information (mean time-dependent AUC = 0.880 versus 0.824). In particular, such improvement was achieved with only one additional image feature, suggesting the importance of using multi-modality data in risk analysis. The laboratory testing results in this study showed that, at the time of admission, the severe group patients already presented a sign of function impairment in organs such as liver (eg: lactate dehydrogenase and prealbumin), heart (eg: multiple types of myocardial enzymes including troponin I, N-terminal brain natriuretic peptide, creatine kinase myocardial band, mitochondrial-Aspartate transaminase and Aspartate transaminase) and kidney (eg: glomerular filtration rate, cystatin C and β2-Microglobulin). The signal of abnormality in blood oxygen also emerges, as PaO2/FiO2, a ratio used to determine severity onset in this study, showed a significant, although not ideal, difference between the two groups upon admission. Consistent with previous studies2,5,6,14,15, the severe group of this cohort demonstrated, at an early stage, a substantial increase in inflammatory factors such as C-reactive protein and interleukin-6, and dysfunction of coagulation. Incorporating laboratory biomarkers tested at an early stage can also significantly improve risk prediction performance as compared to the best model without considering them (mean AUC = 0.884 versus 0.813).

We are still left with a few possible extensions. First, the laboratory testing data had not been integrated into the CT image-based model because blood sample was not collected along with imaging in the center. A more complicated statistical model is required to account for the error caused by time difference inherent in input data. Second, the occurrence of severity may depend on other factors such as treatment, viral load or genetic factors. Our model can include such additional covariates. However, the availability of well-processed data on these factors is scarce, and we have shown that the current model can have reasonable prediction performance even without considering them. Third, because of limited sample size of patients progressed to severity, we only evaluated model prediction performance by cross-validation. Further validation needs to be conducted on external datasets. Fourth, we restricted our analysis of the severe group up to the severity onset. Factors related to the recovery from severe conditions have not been evaluated.

Finally, this study does not include young patients (age<18). To our knowledge, this is the first attempt to estimate the real-time occurrence risk of severity from a cohort featured with a significant amount of imported cases. We believe that the results from this study will be helpful to medical practitioners as they consider how to better manage the care of COVID-19 pneumonia patients upon admission.

## Data Availability

The data can be available from corresponding authors upon reasonable request

## Acknowledgement

This work is funded by Shenzhen Science and Technology Innovation Commission (JSGG20200519160750001) and Sanming Project of Medicine in Shenzhen (SZSM201812058), China.

## Notes

### Competing Interest Statement

The authors have declared no competing interest.

### Funding Statement

This work is partially funded by the Shenzhen Science and Technology Innovation Commission, grant No. JSGG20200519160750001, and Sanming Project of Medicine in Shenzhen, grant No. SZSM201812058.

### Summary of Updates

a new grant number is added as requested by the funding committee.

